# Which brain injury metrics are suitable for supporting sports head injury assessment? A multi-sport brain strain evaluation

**DOI:** 10.64898/2026.06.30.26356946

**Authors:** Emily Yik Kwan Chan, Eleni Koumantou, Lucas Low, Isaac Siy, Chris M. Jones, Kieran Austin, Mike Loosemore, Stuart J. McDonald, Mazdak Ghajari

## Abstract

**Objective:** To identify brain injury metrics suitable for supporting sports head injury assessment by evaluating their association with brain tissue strain and consistency across sports.

**Methods:** Head kinematics from 3,139 impacts in boxing, mixed martial arts, and rugby matches were recorded using instrumented mouthguards and used to calculate nine brain injury metrics. Impacts were simulated using an anatomically detailed finite element brain model to estimate peak 95^th^-percentile maximum principal strain (MPS) in brain and brainstem, a measure of tissue deformation associated with long-term pathology. Sport-specific ordinary least squares models estimated *x_E_*, the metric value equivalent to a reference MPS of 0.21. Metric-MPS correlations and *x_E_* uncertainties were quantified using 5000 bootstrap resamples. Cross-sport consistency was assessed using the coefficient of variation (CV) of sport-specific median *x_E_* values, and uncertainty using the normalised confidence interval size (NCIS).

**Results:** XGB, an extreme gradient boosting strain-prediction model, showed the strongest and most consistent correlations with whole-brain (*r*=0.924–0.974) and brainstem MPS (*r*=0.887–0.954) across all sports. PRV, BrIC and UBrIC also correlated strongly with whole-brain (*r*=0.724–0.930) and brainstem MPS (*r*=0.739–0.900), whereas HIC15 and HARM showed weaker correlation with MPS, particularly in rugby. XGB showed the lowest cross-sport variability (CV=0.034) and uncertainty (median NCIS=0.056). HARM, DAMAGE and HIC15 showed the greatest sport dependence (CV=0.575–0.588) and uncertainty (median NCIS=0.331–0.791).

**Conclusions:** XGB, BrIC, and UBrIC demonstrated the strongest associations with brain tissue strain and the greatest consistency across sports. This study provides a biomechanically informed framework for selecting suitable metrics for sports HIA protocols.

## Introduction

Rapid head motion (kinematics) in sports can cause large brain tissue deformations, leading to traumatic brain injury (TBI). To identify head impacts that produce brain injuries, several brain injury metrics, such as peak linear acceleration (PLA), peak rotational acceleration (PRA), peak rotational velocity (PRV), the head injury criterion (HIC), and the brain injury criterion (BrIC), have been used along with thresholds [1, 2]. For example, a study of collegiate American football proposed that PRA = 6,383 rad/s^2^ and PRV = 28.3 rad/s correspond to an estimated 50% concussion risk [1]. The increased use of instrumented mouthguards (iMGs) in sports has made large-scale, on-field kinematics measurements feasible [3], making some metrics available in real-time to support pitch-side head injury assessment (HIA). For instance, World Rugby uses iMG-derived PLA ≥ 65 g and PRA ≥ 4,500 rad/s^2^ for women and PLA ≥ 75 g and PRA ≥ 4,500 rad/s^2^ for men as assisting triggers for HIA referral [4, 5].

Although threshold values of brain injury metrics have been developed from particular sensor systems, populations and impact environments, they are often used outside their original development context. For instance, concussion-associated thresholds PLA = 95 g and PRA = 5,500 rad/s^2^ obtained from American football data [6] were applied in non-helmeted junior rugby union to classify injurious impacts [7], and subsequently they were carried forward in rugby union studies [8], despite the differences between these sports. Such cross-sport applications may misrepresent injury risk because gameplay, athlete posture, protective equipment, and contact rules influence not only peak values but also duration, time-to-peak, and dominant direction of head kinematics [9, 10, 11], all of which influence brain tissue deformation and injury. In practical terms, this suggests that head-impact monitoring frameworks should not simply adopt a single metric across sports, but they should justify metric choice according to the intended biomechanical interpretation and sport-specific impact profile.

Finite element (FE) brain models, which are biomechanical computational models that incorporate brain anatomical details and nonlinear properties, allow us to translate head kinematics into brain tissue deformation, commonly quantified as maximum principal strain (MPS) [12]. The FE-derived MPS is a sport-agnostic biomechanical endpoint suitable for evaluating brain injury metrics and their thresholds across different sports. Clinical endpoints such as diagnosed concussion or HIA referral can be influenced by symptom reporting, medical resources, and sport-specific assessment protocols, and concussion itself lacks pathological definition [13]. In contrast, MPS provides an objective and mechanistically interpretable reference for comparing metrics across sports contexts. This approach is supported by evidence associating FE-estimated MPS with the location of structural damage [15, 16], neurodegeneration [17], and loss of consciousness [18]. In addition, growing evidence suggests that long-term neuropathological effects of sports head impacts may be related to brain biomechanical exposure than to diagnosed concussion history [14]. Therefore, anchoring injury metric evaluation to MPS allows us to assess how well each metric captures brain-relevant mechanical loading, while recognising that clinical outcomes remain essential for diagnosis, management, and eventual clinical validation.

The aim of this study is to identify which brain injury metrics are more suitable for supporting HIA in different sports by using FE-derived MPS as the endpoint. We evaluated nine candidate metrics using the following criteria: (1) correlation with MPS, sport dependence, assessed by whether the metric value required to reach the same MPS value differs between sports, and (3) pitch-side availability, reflecting whether a metric can be used for in-game injury assessment.

## Methods

### Head kinematics data

We used head kinematics from 3,139 head impacts across four sport datasets in male competitions (Figure 1A): (1) 558 impacts collected during a professional boxing competition, (2) 485 head impacts from amateur mixed martial arts athletes (MMA-A), (3) 395 head impacts from a professional MMA event (MMA-P), and (4) 1,701 head impacts from multiple Rugby Football Union (RFU) games from a professional team. The head kinematics data were collected using the Protecht iMGs (Sports and Wellbeing Analytics, Swansea, UK), which has been validated in previous studies [19, 20]. All impacts were video verified and underwent the mouthguard’s standard data processing [19, 21].

**Figure 1.**
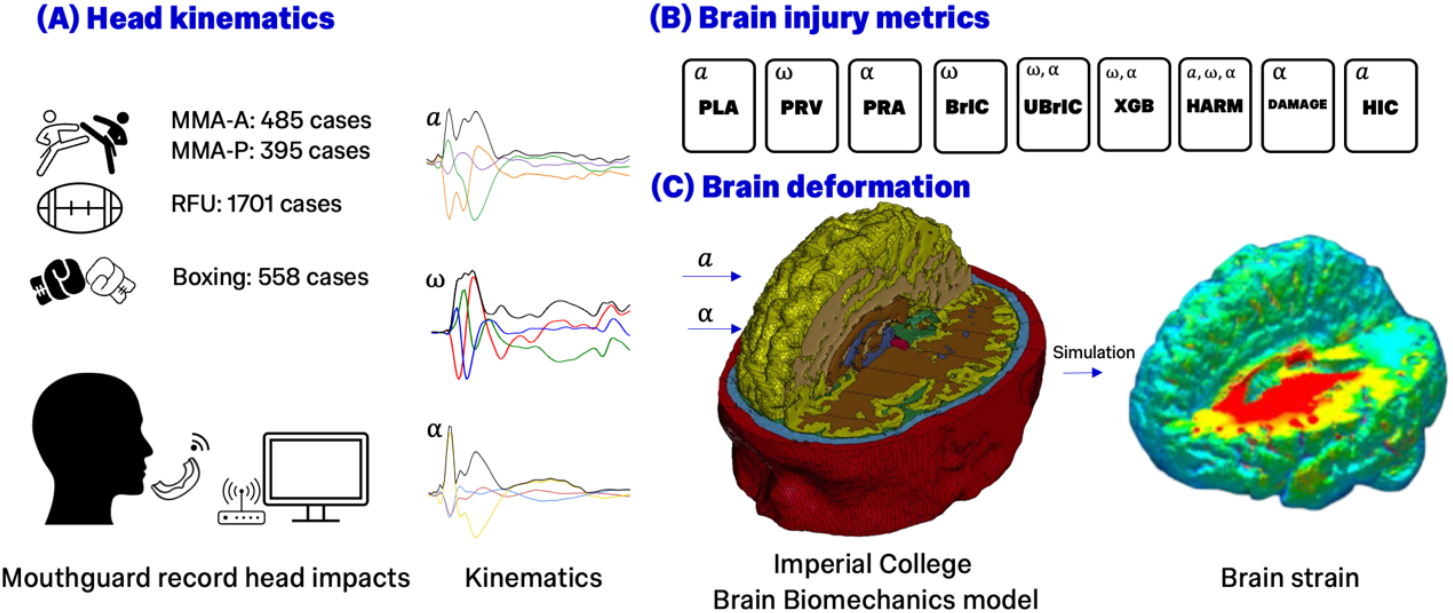
Methods. (A) Head impact kinematics were collected using instrumented mouthguards from multiple sports, (B) values of widely used injury metrics were calculated for each impact, and (C) brain strains produced by each impact were estimated.

### Brain strain prediction

The anatomically detailed Imperial College (IC) FE brain model was used to predict brain strain [17] (Figure 1C). This model has been validated against cadaveric rotational tests by comparing simulated brain displacements with sonomicrometry measurements [22], and it has been used to predict the location of chronic traumatic encephalopathy (CTE) pathology and provide a biomechanical explanation for loss of consciousness [17, 18]. To simulate each impact, 103 ms of head linear and rotational accelerations were applied to the brain model. The LS-DYNA explicit solver was used for simulations (version 971, R9.1.0; Ansys, Canonsburg, PA, USA).

For every element of the brain model, the peak value of the maximum principal Green-Lagrange strain during the simulation was computed. The resulting strains were exported to neuroimaging NIfTI files. The 95th percentile maximum principal strain (MPS) was extracted for the whole brain (WB) as a measure of global tissue deformation, and for the brainstem, a region associated with loss of consciousness [18].

### Brain injury metrics

We evaluated nine brain injury metrics for each impact (Figure 1B and Table 1). We calculated the resultant peak linear acceleration (PLA), peak rotational acceleration (PRA), and peak rotational velocity (PRV), which are widely used in sports [4, 6]. The Head Injury Criterion (HIC) was also included, which uses the resultant linear acceleration and was originally developed for automotive applications [23] but has been used in sports [24, 25].

**Table 1.**
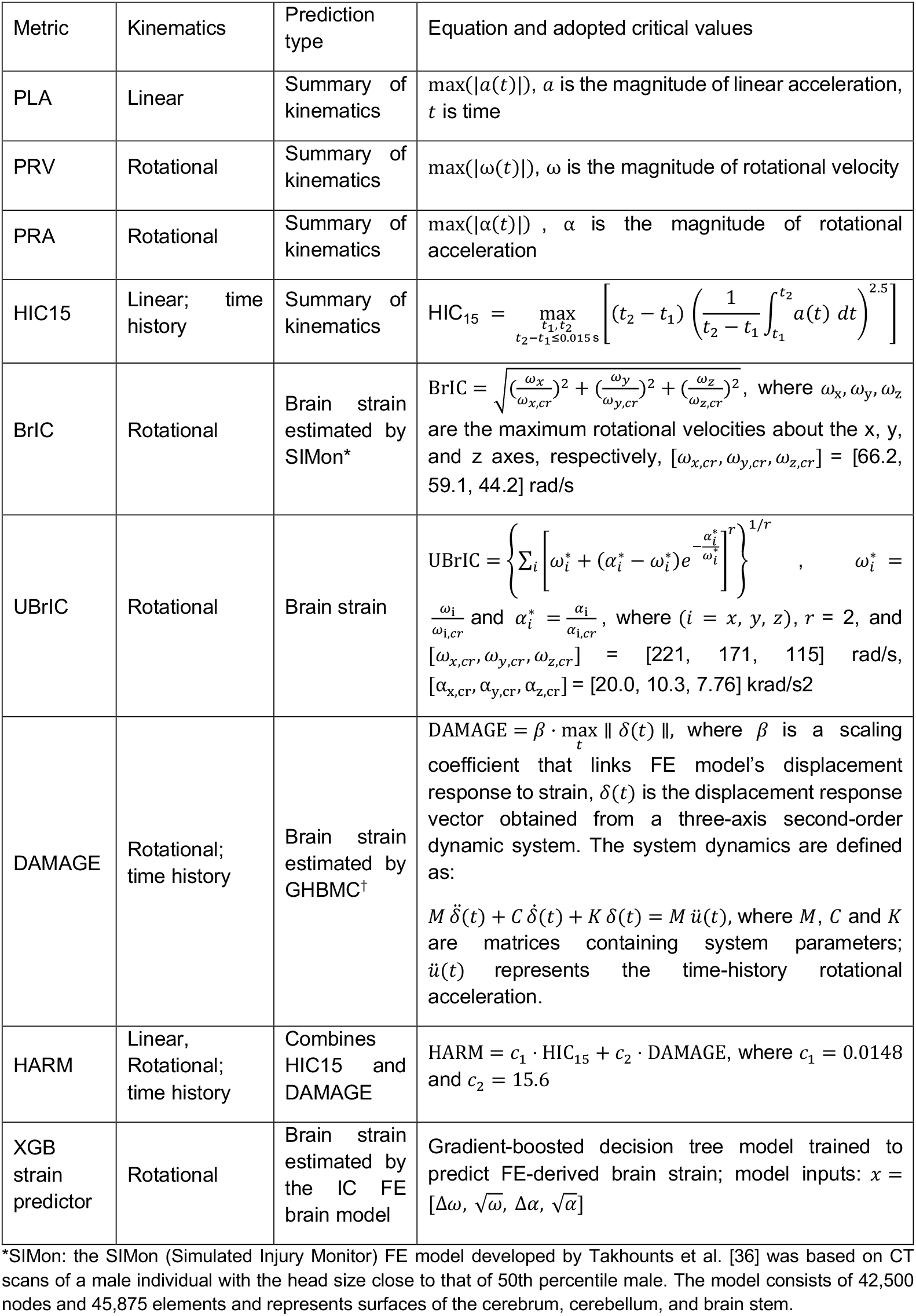
Summary of brain injury metrics.

The Brain Injury Criterion (BrIC) was developed to predict brain strain estimated by the SIMon^*^ brain model, with directionally dependent critical values of PRV calibrated to predict diffuse axonal injury [2]. BrIC has been used in iMG validation and cycling helmet safety ranking [21, 26]. A modified version of BrIC is the Universal Brain Injury Criterion (UBrIC), which combines directional peak rotational velocities and accelerations, and was validated against the SIMon and GHBMC^†^ brain models [27].

The Diffuse Axonal Multi-Axis General Evaluation (DAMAGE) metric has also been developed to predict brain strain using three components of rotational acceleration time histories [28]. The model parameters, including mass, stiffness, and damping, were tuned to match the GHBMC brain model’s strain predictions. DAMAGE has been used in multiple sport-related studies [29, 30].

The Head Acceleration Response Metric (HARM) linearly combines HIC15 and DAMAGE to assess head impact severity and is used in National Football League (NFL) helmet performance scoring [31, 32]. We adopted weighting constants determined through reconstructions of American football impacts involving concussed and non-concussed players [33, 34].

We also used a new model developed via the eXtreme Gradient Boosting (XGB) machine learning algorithm [35]. The XGB model predicts brain strain estimated by the IC FE brain model using two features from resultant rotational velocity and acceleration time histories: the maximum and its square root. We updated the model’s weightings by retraining it on 20% of the sport dataset used in this study.

### Statistical analysis

We compared MPS across sport datasets using Kruskal–Wallis tests separately for whole‐brain and brainstem followed by Mann–Whitney U tests for post‐hoc pairwise comparisons due to the skewed distribution of strain data.

To evaluate the suitability of each metric for supporting sports HIA, we assessed three key properties: (i) pitch-side availability, (ii) association with brain strain, and (iii) cross-sport consistency of the estimated metric values required to reach the same MPS level, together with the uncertainty of these estimates.

Pitch-side availability was inferred from the metrics input: metrics that require full kinematic time histories might be less readily pitch-side available in current iMG workflows, as contemporary iMGs can only transmit a few kinematic features in real time rather than full kinematic time histories.

To quantify metric-MPS association in each sport, we calculated the Pearson correlation coefficient (*r*) using 5000 bootstrap resamples with replacement and reported the mean ± standard deviation (SD) of the resulting *r* values.

We fitted a univariate ordinary least squares (OLS) regression model with MPS as the outcome and the metric as the predictor for each metric within each sport dataset. OLS was used because HIA metrics are typically applied in practice as single cut-off values, rather than as non-linear calibration models. From each fitted model, we derived *x*_*E*_, the metric value at whole-brain MPS reference level of 0.21. This reference strain level has been previously associated with tissue injury [37]. It is also consistent with a parallel biomarker analysis from our group, where segmented regression identified a change point near 0.21 in the relationship between FE-estimated whole-brain MPS and plasma glial fibrillary acidic protein (GFAP), a blood biomarker used to detect damage and inflammation in the central nervous system [38]. We used 5000 within-sport bootstrap iterations with replacement to estimate *x*_*E*_ for each metric-sport combination.

To assess whether a given metric produced similar *x*_*E*_ values across sports, we calculated the coefficient of variation (CV). For each metric, CV was calculated across the four sport-specific median *x*_*E*_ values. Metrics with lower CV values indicate that the *x*_*E*_ values are more consistent across sports. To quantify uncertainty in the derived *x*_*E*_ values, we determined the 95% confidence interval (CI) for each metric-sport combination and calculated the normalised confidence interval size (NCIS), defined as the width of the 95% CI divided by the absolute median *x*_*E*_. Lower NCIS values indicate more precise *x*_*E*_ estimates.

## Results

### Brain strain distribution across sports

The brain strain distribution was different across sports (Figure 2). The voxel-wise mean strain maps illustrate the average spatial pattern of tissue deformation within each sport, whereas the boxplots summarise the distribution of impact-level MPS values used for statistical analysis. The median and interquartile range (IQR) of whole-brain MPS was 0.077 (0.06) for RFU, 0.066 (0.055) for amateur MMA, 0.099 (0.095) for professional MMA, and 0.127 (0.104) for boxing. The Kruskal–Wallis test indicated significant between-sport differences in MPS for both the whole brain (H = 377.76, p < 0.001) and brainstem (H = 372.87, p < 0.001). Pairwise Mann–Whitney U tests further confirmed that MPS in both regions differed significantly between all pairs (all p < 0.001).

**Figure 2.**
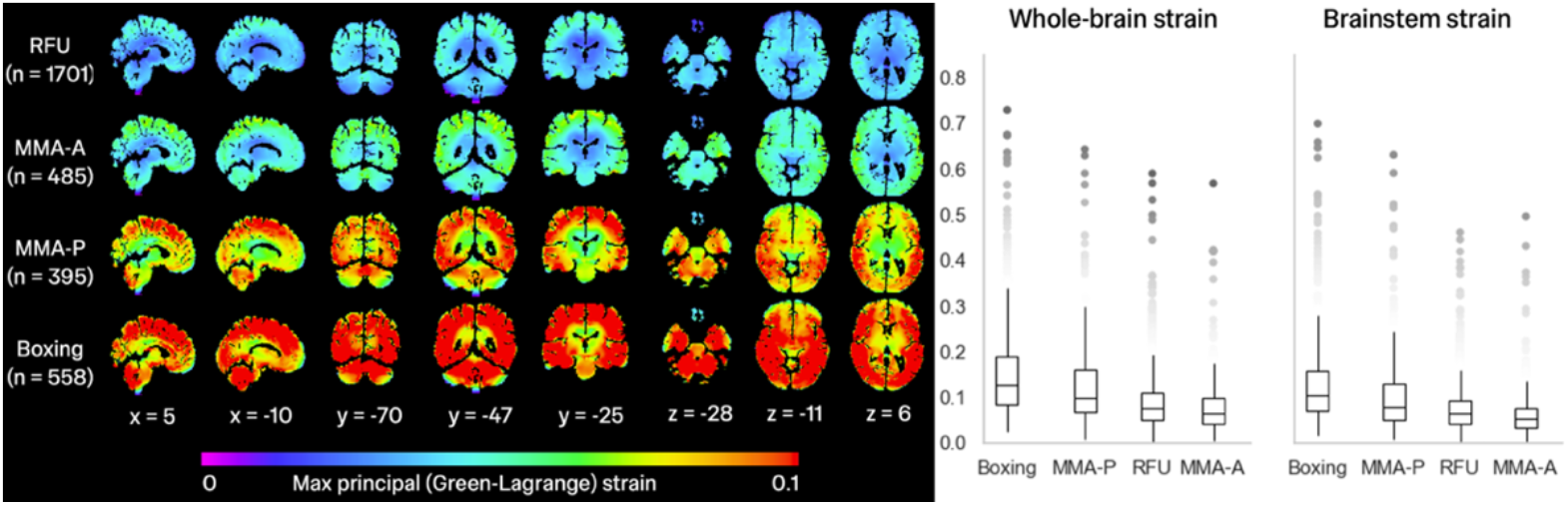
Strain distribution across sport datasets. Left panel: voxel-wise mean maximum principal strain (MPS) maps calculated by averaging strain at each voxel across all impacts within each sport dataset, illustrating the average spatial pattern of tissue deformation. Columns show representative sagittal (x), coronal (y), and axial (z) slices, with coordinates provided in MNI space. Right panel: distributions of impact-level 95th-percentile MPS in the whole brain and brainstem for each sport dataset, which were used for the statistical analyses.

### The correlation between injury metrics and brain strain

Strongest correlations (*r* > 0.9) were found for XGB strain predictor across all sports, for PRV and PRA across boxing and MMA, for PLA in boxing, and for BrIC in MMA (Figure 3a). HIC15, DAMAGE, and HARM showed generally weaker correlations (*r* < 0.8) across all sports. Rugby showed the weakest correlation across all metrics. A similar pattern of correlations was found between the metrics and brainstem MPS (Figure 3b).

**Figure 3.**
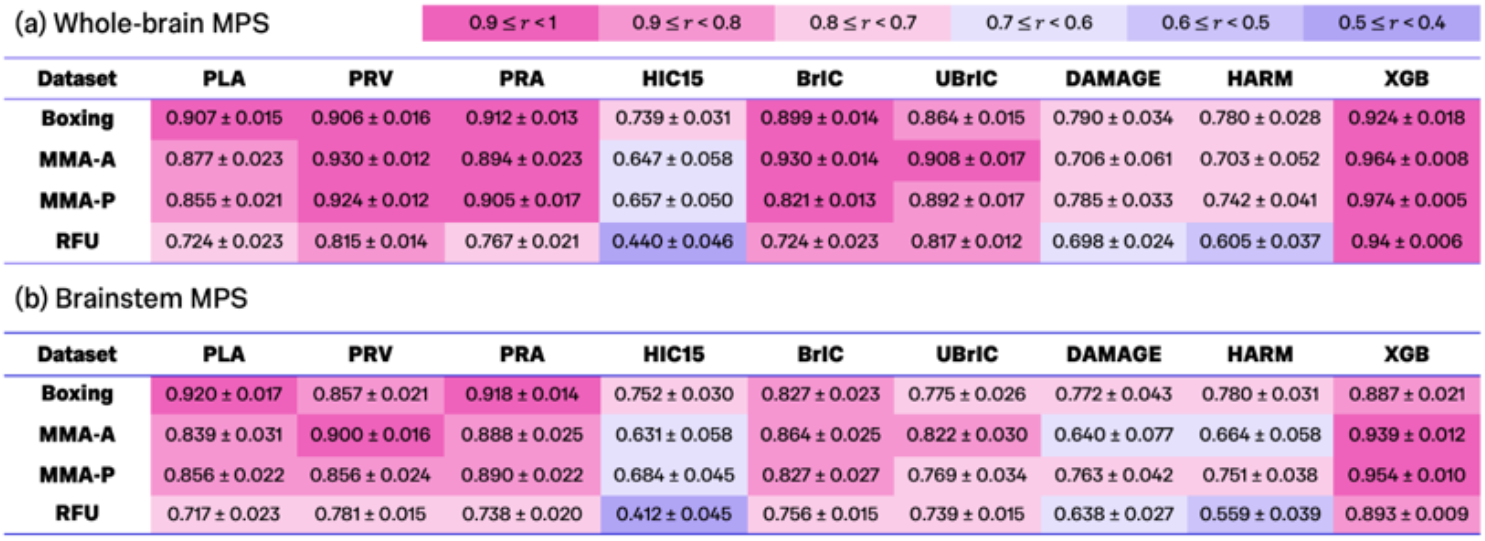
Bootstrap mean Pearson correlation coefficients (r) mean ± standard deviation (SD) between each kinematic metric and FE-estimated MPS in **(a)** the whole brain and **(b)** the brainstem across the four sport datasets. Cells are colour-coded by the mean r to show the strength of correlation.

### Sport dependence of injury metrics

Bootstrap distributions of the reference-equivalent metric value *x*_*E*_ showed varying degrees of spread within sports and separation between sports across metrics (Figure 4a). The coefficient of variation (CV) calculated across the sport-specific median *x*_*E*_ values is shown in Figure 4b. The XGB strain predictor showed the lowest CV (0.034), indicating the greatest sport agnosticity, followed by rotational-based metrics UBrIC (0.102), BrIC (0.126), and PRV (0.139). PLA and PRA showed intermediate cross-sport variation, with CV values of 0.329 and 0.348, respectively. In contrast, HARM (0.575), HIC15 (0.579), and DAMAGE (0.588) showed the highest CV values, indicating greater sport dependence, with substantially different metric values required to reach the same whole-brain strain level across sports, as shown in Figure 4a.

**Figure 4.**
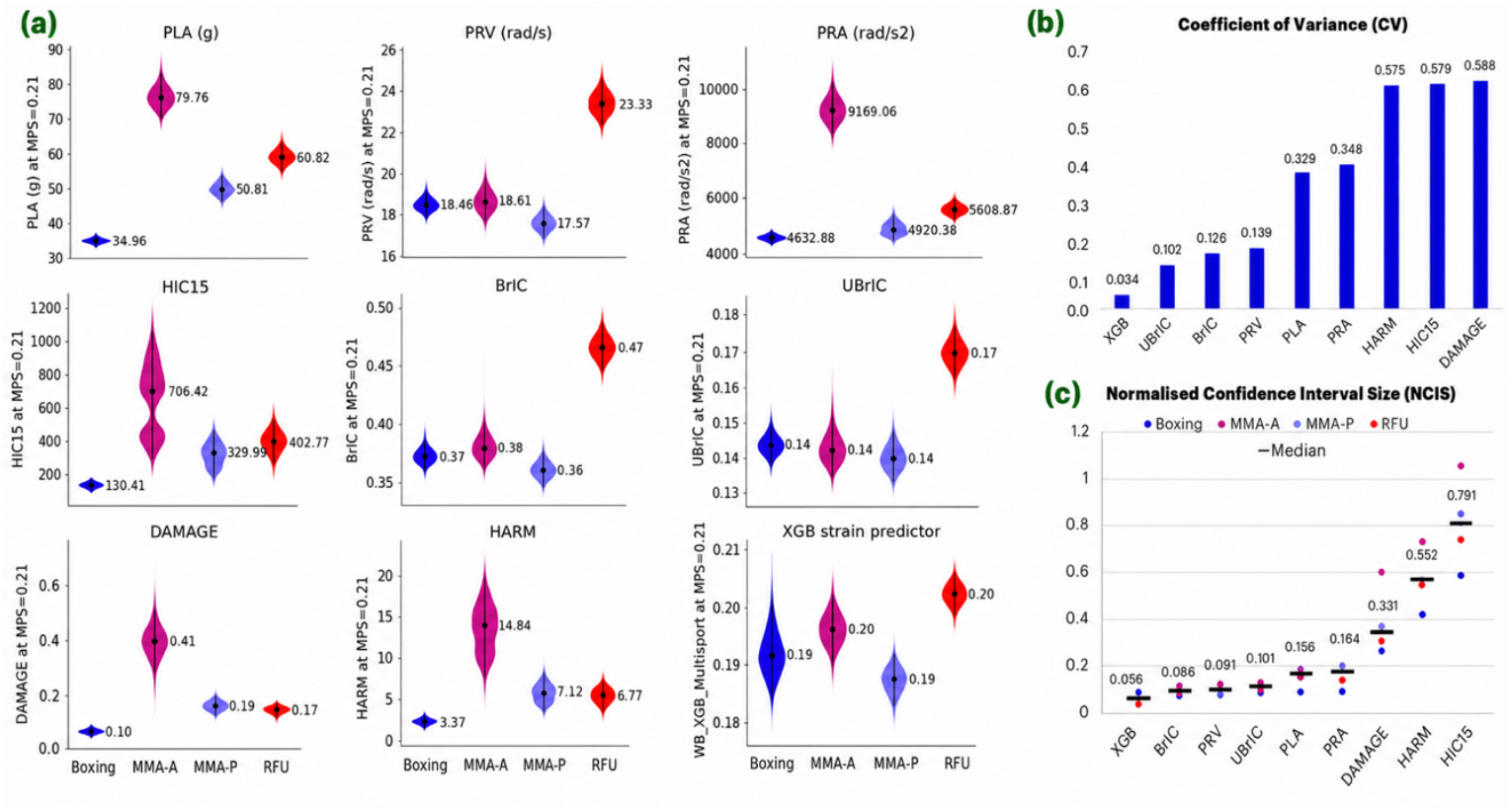
Sport-specific reference-equivalent metric values x_E_ at whole-brain MPS=0.21 reference. (a) Bootstrap distributions of the reference-equivalent metric value (x_E_) for each metric and sport. Black points and lines indicate the bootstrap median and 95% CI, respectively. (b) Cross-sport coefficient of variation (CV) of sport-median x_E_ values, summarising sport dependence of each metric. Lower CV indicates greater sport agnosticity. (c) Normalised confidence interval size (NCIS), calculated as CI width divided by the median x_E_, summarising the precision of x_E_ estimation within each sport; horizontal markers indicate the median NCIS across sports for each metric. Lower NCIS indicates less uncertainty.

The median normalised confidence interval size (NCIS) for each metric across the four sports is shown Figure 4c. The XGB strain predictor had the lowest median NCIS (0.056), followed by BrIC (0.086), PRV (0.091), UBrIC (0.101), PLA (0.156), and PRA (0.164), whereas HARM (0.331), DAMAGE (0.552), and HIC15 (0.791) had higher median NCIS values, indicating less precise estimation of the reference-equivalent metric value, *x*_*E*_.

Detailed bootstrap summaries for *x*_*E*_ (median, IQR, 95% CI and NCIS for each sport and metric) are provided in Supplementary Table S1. An equivalent analysis for brainstem is shown in Supplementary Figure S1 and Supplementary Table S2.

## Discussion

This study showed that brain injury metrics differ not only in their association with FE-estimated brain strain, but also in the extent to which metric values correspond to the same level of brain strain across sports. This finding has important implications for head injury assessment (HIA), as threshold values are often adopted from studies conducted in different sports populations. Our results suggest that such thresholds may not be directly transferable, with some metrics exhibiting substantial sport-specific variation in the metric value associated with a given level of tissue strain. These observations were enabled by a large multi-sport dataset collected using a single instrumented mouthguard (iMG) system and processed through a consistent pipeline, thus minimising potential iMG- and processing-related confounding. Among the nine metrics evaluated, the XGB strain predictor, BrIC and UBrIC showed the most favourable combination of strong association with brain strain, low sport dependence and low uncertainty in the reference-equivalent metric value. These metrics therefore appear to provide the most robust basis for supporting HIA decision-making across different sporting contexts.

### Transferability of injury thresholds between sports

A key finding of this study is that some injury metrics correspond more consistently than others to a given level of brain strain across sports. The XGB strain predictor showed the lowest cross-sport variability, followed by UBrIC, BrIC and PRV, whereas HIC15, HARM and DAMAGE showed the greatest sport dependence and uncertainty. These findings suggest that thresholds based on XGB, BrIC or UBrIC are more likely to correspond to comparable tissue-level loading across boxing, MMA and rugby, whereas thresholds based on HIC15, HARM or DAMAGE may require recalibration for each sport.

This consistency is particularly important for supporting HIA referral decisions. If the metric value associated with a given level of brain strain differs substantially between sports, then applying a common threshold may result in different levels of tissue deformation across sporting contexts. Consequently, injury thresholds derived in one sport may not be directly transferable to another.

This has important practical implications because much of the existing evidence underpinning head impact thresholds has been developed within specific sporting contexts, particularly American football and rugby [1, 39]. However, sports differ in impact mechanism, contact location, athlete posture, anticipation, protective equipment and the relative contribution of linear and rotational motion. For example, striking sports, such as boxing and MMA, expose athletes to different impact profiles than rugby and American football [39, 40]. These differences are biomechanically important because impact direction, loading duration and the relative contributions of linear and rotational motion influence how external head kinematics translate into brain tissue deformation [9, 10, 11]. In addition, athlete posture, anticipation and neck muscle activation may further modify the kinematic response of the head during impact [41]. Together, these factors suggest that a metric that performs well in one sport may not necessarily retain the same biomechanical interpretation in another.

### Biomechanical validity of candidate metrics

We also found that metrics derived from rotational head kinematics or explicitly informed by brain strain generally showed stronger associations with FE-estimated brain strain than metrics based mainly on linear acceleration or combined legacy criteria. The XGB strain predictor showed the strongest correlations with both whole-brain and brainstem MPS across all sports. PRV, BrIC and UBrIC also showed strong correlations, whereas HIC15, DAMAGE and HARM showed weaker correlations, particularly in rugby. These findings are consistent with previous work showing that rotational kinematics are more strongly related to brain strain than linear acceleration [30, 42, 43].

The strong performance of rotational and strain-informed metrics has important implications for HIA. Unlike metrics that primarily summarise external head motion, these metrics are designed to better reflect the mechanical loading experienced by brain tissue. This distinction is important because FE-estimated brain strain has been associated with acute symptoms, neuroimaging abnormalities and other objective markers of injury [15, 16, 17, 18]. This supports the use of MPS as a biologically meaningful and mechanistically relevant reference for evaluating injury metrics across sports, while recognising that clinical assessment remains essential for diagnosis and management.

However, our results also indicate that being strain-informed does not guarantee transferability across sports or impact contexts. Although DAMAGE and HARM were developed using FE-model-derived brain strains, they showed weaker and less consistent relationships with IC-model-derived strain in the present multi-sport dataset. Both metrics were developed and calibrated using different FE models and impact datasets, including automotive and American football exposures [28, 31, 32]. Their reduced performance may therefore reflect differences in the FE models, impact characteristics and calibration datasets. These findings suggest that strain-informed metrics may still require calibration when applied to new sporting environments, highlighting the importance of evaluating both biomechanical relevance and cross-sport robustness when selecting metrics for HIA applications.

### Role of the XGB strain predictor

The strong performance of the XGB strain predictor should be interpreted in context. XGB is a reduced-order surrogate of the IC FE brain model and was designed to predict FE-derived strain from kinematic features [35]. Its high correlation with IC FE-derived MPS is therefore expected and should not be interpreted as independent clinical validation. Instead, its value in this comparison is to show how closely a near-real-time brain surrogate can reproduce the reference FE model across different sports. This approach is consistent with previous development and evaluation of FE-informed injury metrics, such as BrIC, UBrIC and DAMAGE, where kinematic metrics were derived from, or evaluated against, FE-model strain responses [2, 27, 42]

### Operational feasibility for pitch-side HIA

For a metric to support HIA, it must also be feasible in pitch-side workflows. Metrics such as PLA, PRV, PRA, BrIC, UBrIC and the XGB strain predictor can be computed from summary kinematic features and are therefore readily compatible with real-time or near-real-time iMG systems. Metrics such as HIC15, DAMAGE and HARM require fuller waveform information or additional processing and may therefore depend more strongly on the technical capabilities of the iMG system and sideline infrastructure. This does not preclude their near-real-time use, particularly where waveform processing can be performed on-device or where full kinematic signals can be transmitted reliably to pitch side. However, such workflows may require greater on-device computational capacity, more robust data transmission, or additional sideline processing infrastructure, which could affect cost, scalability and suitability for community or grassroots implementation. Therefore, when considering pitch-side HIA, metric selection should account not only for biomechanical performance, but also for the practical requirements of real-world deployment.

### Limitations

We analysed head kinematics recorded by the Protecht iMG. This reduced device-related confounding, but the findings may not directly generalise to other iMGs because sampling rate, triggering thresholds, recording windows and signal processing can differ between systems. It should also be noted that the observed sport differences may partly reflect the sampled impacts within each sport, rather than intrinsic properties of the sports themselves. Larger and more representative datasets are therefore needed to confirm the generalisability of these findings. In addition, we assessed single-impact brain deformation only and did not examine cumulative effects of repetitive impacts, as information on inter-impact intervals was unavailable. Consequently, the estimated strain should not be interpreted as the safety or the injury risk of each sport. Finally, this study used FE-estimated MPS as a mechanistic reference [12, 17, 18] rather than clinical outcome data. Future studies should combine iMG kinematics, FE modelling, and clinical or biological measures of injury, such as blood-based biomarkers, to further validate candidate HIA metrics [38, 44, 45].

## Conclusions and Implications

This study shows that brain injury metrics intended to support sport HIA should be evaluated not only by their association with brain strain, but also by their cross-sport consistency and pitch-side feasibility. We found that both metric–strain associations and reference-equivalent metric values varied across sports, indicating that thresholds developed in one sport may not necessarily correspond to the same tissue-level loading in another. Among the candidate metrics, XGB, BrIC and UBrIC showed the best overall combination of near-real-time feasibility, strong association with FE-estimated brain strain, and lower sport dependence. These findings support the use of strain-informed metrics to complement conventional kinematic metrics and suggest that sport-specific recalibration may be needed when sport-dependent metrics are used for HIA thresholds.

## Acknowledgements

EYKC acknowledges the postgraduate funding support by Sports & Wellbeing Analytics. MG acknowledges the support of Royal Academy of Engineering Senior Research Fellowship (RCSRF2324-17-19). The authors also acknowledge the wider team at Sports & Wellbeing Analytics for their assistance with the data collection. The results of this study are solely produced by the authors and do not constitute the endorsement of any party.

## Statements and Declarations

### Contributors

EYKC: conception, design, simulation, data analysis, manuscript writing. EK, IS: simulation. LL: manuscript editing. CJ, KA, ML: data collection and curation, manuscript editing. MG: conception, design, manuscript editing, supervision.

### Data availability

All injury metrics data relevant to this study are included in the article or supplementary. The instrumented mouthguard data are not publicly available due to the sensitive nature of the data and the privacy of the participants.

### Ethics approval

Sports & Wellbeing Analytics collects head impact data with consent from participating athletes. The collected data are shared with Imperial College London for research under the ethics approval of the Imperial College Research Governance and Integrity Team (ICREC-6439674).

