## Supplementary materials for "Which brain injury metrics are suitable for supporting sports head injury assessment? A multi-sport brain strain evaluation"

### S1. Brainstem reference-equivalent analysis using a 0.19 MPS reference

In the main analysis, reference-equivalent metric values were estimated using whole-brain MPS at a reference value of 0.21. As a supplementary regional analysis, we repeated the same sport-specific OLS and bootstrap procedure using brainstem MPS. The brainstem reference value was set to 0.19, based on the blood biomarker study by Hickey et al. (under review), which reported a breakpoint near this level for brainstem strain. As for the whole-brain analysis, this value was used as a fixed biomechanical reference level for comparing metrics across sports, rather than as a validated clinical injury threshold.

For each metric and sport, sport-specific OLS models were used to estimate the metric value equivalent to a brainstem MPS of 0.19. Bootstrap resampling was used to quantify uncertainty in these estimates. Cross-sport consistency was summarised using the coefficient of variation (CV) of sport-specific median $x_E$ values, and uncertainty was summarised using the normalised confidence interval size (NCIS), as described in the main Methods.

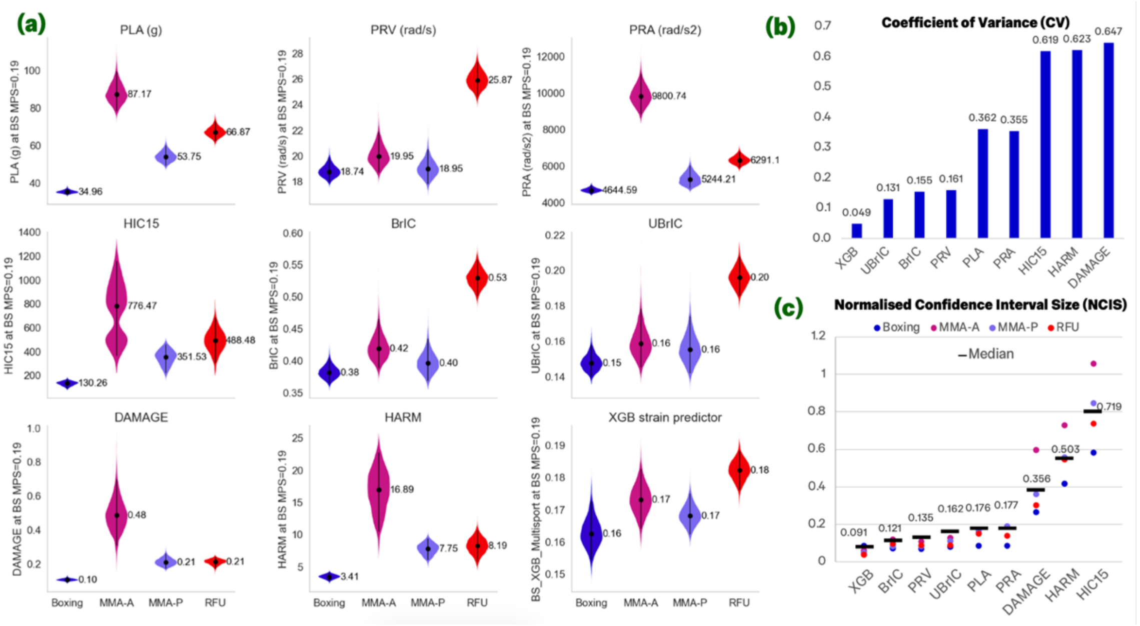


*Supplementary Figure S1 Sport-specific reference-equivalent metric values $x_E$ at brainstem MPS = 0.19 reference. (a) Bootstrap distributions of the reference-equivalent metric value ($x_E$) for each metric and sport. Black points and lines indicate the bootstrap median and 95% CI, respectively. (b) Cross-sport coefficient of variation (CV) of sport-median $x_E$ values, summarising sport dependence of each metric. Lower CV indicates greater sport agnosticity. (c) Normalised confidence interval size (NCIS), calculated as CI width divided by the median $x_E$, summarising the precision of $x_E$ estimation within each sport; horizontal markers indicate the median NCIS across sports for each metric. Lower NCIS indicates less uncertainty.*

The brainstem analysis showed a similar overall pattern to the whole-brain analysis. The XGB strain predictor showed the greatest cross-sport consistency, with the lowest CV across sports (CV = 0.049; Supplementary Table S1). UBrIC also showed relatively low cross-sport variation (CV = 0.131), followed by BrIC (CV = 0.155) and PRV (CV = 0.161). In contrast, DAMAGE (CV = 0.648), HARM (CV = 0.623), and HIC15 (CV = 0.619) showed substantially greater variation in reference-equivalent values across sports.

The XGB strain predictor also showed consistently low uncertainty across sports, with NCIS values ranging from 0.066 to 0.109 (Supplementary Table S2). PRV, BrIC, UBrIC, PLA, and PRA generally showed low-to-moderate uncertainty, whereas HIC15, DAMAGE, and HARM showed larger uncertainty, particularly in MMA-A. Overall, the supplementary brainstem analysis supported the main whole-brain findings, indicating that the XGB strain predictor provided the most consistent reference-equivalent values across sports, while several conventional kinematic and composite metrics showed greater sport dependence.

### S2. Full bootstrap summaries of whole-brain and brainstem reference-equivalent metric values

This section provides the full data of the OLS/bootstrap table for both WB and BS. For each sport and metric, the table reports the bootstrap median reference-equivalent value $x_E$, interquartile range (IQR), percentile 95% confidence interval (2.5th–97.5th percentiles), and the normalised confidence interval size (NCIS), computed as CI width divided by the absolute median $(x_E)$, using 5000 within-sport bootstrap resamples.

*Supplementary Table S1 Bootstrap summary of sport-specific reference-equivalent metric values for the whole brain.*

| Metric | Sport | x_E median | x_E IQR | x_E CI (low) | x_E CI (high) | x_E NCIS |
|---|---|---|---|---|---|---|
| PLA (g) | Boxing | 34.964 | 0.996 | 33.603 | 36.608 | 0.086 |
| | MMA-A | 79.759 | 4.758 | 73.563 | 87.444 | 0.174 |
| | MMA-P | 50.815 | 2.932 | 47.023 | 55.233 | 0.162 |
| | RFU | 60.823 | 3.037 | 56.722 | 65.821 | 0.150 |
| PRV (rad/s) | Boxing | 18.459 | 0.451 | 17.884 | 19.220 | 0.072 |
| | MMA-A | 18.608 | 0.737 | 17.726 | 19.925 | 0.118 |
| | MMA-P | 17.570 | 0.503 | 16.889 | 18.387 | 0.085 |
| | RFU | 23.329 | 0.800 | 22.255 | 24.530 | 0.098 |
| PRA (rad/s2) | Boxing | 4632.880 | 142.749 | 4444.312 | 4853.254 | 0.088 |
| | MMA-A | 9169.065 | 591.427 | 8358.785 | 10097.699 | 0.190 |
| | MMA-P | 4920.380 | 360.650 | 4529.868 | 5467.850 | 0.191 |
| | RFU | 5608.874 | 269.402 | 5255.600 | 6034.854 | 0.139 |

| | | | | | | |
|---|---|---|---|---|---|---|
| HIC15 | Boxing | 130.405 | 29.495 | 90.396 | 166.348 | 0.582 |
| | MMA-A | 706.421 | 387.200 | 316.379 | 1062.284 | 1.056 |
| | MMA-P | 329.928 | 109.873 | 189.292 | 468.373 | 0.846 |
| | RFU | 402.770 | 95.847 | 255.749 | 552.375 | 0.736 |
| BrIC | Boxing | 0.373 | 0.009 | 0.361 | 0.387 | 0.070 |
| | MMA-A | 0.380 | 0.014 | 0.363 | 0.405 | 0.110 |
| | MMA-P | 0.361 | 0.010 | 0.348 | 0.378 | 0.083 |
| | RFU | 0.465 | 0.014 | 0.445 | 0.487 | 0.089 |
| UBrIC | Boxing | 0.143 | 0.004 | 0.138 | 0.150 | 0.081 |
| | MMA-A | 0.142 | 0.006 | 0.134 | 0.153 | 0.129 |
| | MMA-P | 0.139 | 0.005 | 0.132 | 0.148 | 0.111 |
| | RFU | 0.170 | 0.005 | 0.163 | 0.178 | 0.090 |
| DAMAGE | Boxing | 0.101 | 0.009 | 0.088 | 0.114 | 0.265 |
| | MMA-A | 0.408 | 0.078 | 0.284 | 0.527 | 0.596 |
| | MMA-P | 0.187 | 0.024 | 0.155 | 0.222 | 0.361 |
| | RFU | 0.173 | 0.018 | 0.142 | 0.194 | 0.301 |
| HARM | Boxing | 3.373 | 0.511 | 2.666 | 4.072 | 0.417 |
| | MMA-A | 14.840 | 4.550 | 9.382 | 20.183 | 0.728 |
| | MMA-P | 7.119 | 1.596 | 5.243 | 9.206 | 0.557 |
| | RFU | 6.768 | 1.320 | 4.845 | 8.544 | 0.546 |
| XGB strain predictor | Boxing | 0.192 | 0.006 | 0.185 | 0.201 | 0.087 |
| | MMA-A | 0.196 | 0.004 | 0.190 | 0.202 | 0.062 |
| | MMA-P | 0.188 | 0.003 | 0.182 | 0.192 | 0.051 |
| | RFU | 0.202 | 0.003 | 0.198 | 0.206 | 0.039 |

*Supplementary Table S2 Bootstrap summary of sport-specific reference-equivalent metric values for the brainstem.*

| Metric | Sport | x_E median | x_E IQR | x_E CI (low) | x_E CI (high) | x_E NCIS |
|---|---|---|---|---|---|---|
| PLA (g) | Boxing | 34.955 | 1.079 | 33.607 | 36.768 | 0.090 |
| | MMA-A | 87.166 | 7.268 | 78.363 | 99.210 | 0.239 |
| | MMA-P | 53.754 | 3.738 | 49.134 | 59.719 | 0.197 |
| | RFU | 66.865 | 3.416 | 62.231 | 72.538 | 0.154 |
| PRV (rad/s) | Boxing | 18.745 | 0.606 | 17.984 | 19.776 | 0.096 |
| | MMA-A | 19.952 | 1.063 | 18.706 | 21.819 | 0.156 |

| | | | | | | |
|---|---|---|---|---|---|---|
| | MMA-P | 18.948 | 0.892 | 17.802 | 20.447 | 0.140 |
| | RFU | 25.871 | 0.932 | 24.633 | 27.283 | 0.102 |
| PRA (rad/s2) | Boxing | 4644.586 | 147.615 | 4454.030 | 4886.631 | 0.093 |
| | MMA-A | 9800.743 | 751.057 | 8827.856 | 10988.337 | 0.220 |
| | MMA-P | 5244.210 | 464.179 | 4769.334 | 5979.484 | 0.231 |
| | RFU | 6291.148 | 293.343 | 5903.594 | 6745.798 | 0.134 |
| HIC15 | Boxing | 130.259 | 24.966 | 91.322 | 159.793 | 0.526 |
| | MMA-A | 776.465 | 411.909 | 360.861 | 1152.826 | 1.020 |
| | MMA-P | 351.527 | 88.281 | 218.721 | 454.816 | 0.672 |
| | RFU | 488.484 | 125.737 | 302.116 | 676.364 | 0.766 |
| BrIC | Boxing | 0.381 | 0.013 | 0.364 | 0.403 | 0.101 |
| | MMA-A | 0.418 | 0.026 | 0.388 | 0.464 | 0.183 |
| | MMA-P | 0.395 | 0.021 | 0.368 | 0.433 | 0.165 |
| | RFU | 0.529 | 0.019 | 0.502 | 0.557 | 0.105 |
| UBrIC | Boxing | 0.147 | 0.006 | 0.140 | 0.157 | 0.119 |
| | MMA-A | 0.158 | 0.011 | 0.145 | 0.178 | 0.209 |
| | MMA-P | 0.155 | 0.011 | 0.142 | 0.174 | 0.205 |
| | RFU | 0.196 | 0.008 | 0.185 | 0.207 | 0.111 |
| DAMAGE | Boxing | 0.103 | 0.009 | 0.090 | 0.118 | 0.269 |
| | MMA-A | 0.483 | 0.125 | 0.308 | 0.692 | 0.797 |
| | MMA-P | 0.208 | 0.030 | 0.172 | 0.256 | 0.405 |
| | RFU | 0.210 | 0.022 | 0.171 | 0.236 | 0.308 |
| HARM | Boxing | 3.409 | 0.444 | 2.725 | 4.001 | 0.374 |
| | MMA-A | 16.887 | 4.774 | 10.432 | 22.654 | 0.724 |
| | MMA-P | 7.750 | 1.265 | 5.995 | 9.305 | 0.427 |
| | RFU | 8.194 | 1.710 | 5.773 | 10.514 | 0.579 |
| XGB strain predictor | Boxing | 0.162 | 0.006 | 0.155 | 0.172 | 0.109 |
| | MMA-A | 0.173 | 0.006 | 0.164 | 0.183 | 0.106 |
| | MMA-P | 0.168 | 0.004 | 0.162 | 0.175 | 0.075 |
| | RFU | 0.182 | 0.004 | 0.176 | 0.188 | 0.066 |
